# TRANSGENERATIONAL EFFECT OF EARLY CHILDHOOD FAMINE EXPOSURE IN THE COHORT OF LENINGRAD SIEGE SURVIVORS’ OFFSPRING

**DOI:** 10.1101/2022.11.04.22281954

**Authors:** Kristina Tolkunova, Dmitrii Usoltsev, Ekaterina Moguchaia, Maria Boyarinova, Ekaterina Kolesova, Anastasia Erina, Trudy Voortman, Elena Vasilyeva, Anna Kostareva, Evgeny Shlyakhto, Alexandra Konradi, Oxana Rotar, Mykyta Artomov

**Author notes:** contributed equally (Kristina Tolkunova, Dmitrii Usoltsev).

## Abstract

**Introduction:** Environmental exposure during early life development can affect disease risk in late-life period. There is little data on the transmission of phenotypic features from famine-exposed individuals to the next generations.

**Objective:** The purpose of our study was to investigate the association of parental starvation in the perinatal period and the period of early childhood with the phenotypic features observed in two generations of descendants of Leningrad Siege survivors (DLSS).

**Design and methods:** We examined 54 children and 30 grandchildren of 58 besieged Leningrad residents who suffered from starvation in early childhood and prenatal age during the Second World War. Controls from the population-based national epidemiological ESSE-RF study (n=175) were matched on sex, age and body mass index (BMI). Anthropometry, blood pressure (BP) measurement, and blood tests were performed for both groups. Sociodemographic and lifestyle information was collected via questionnaires. Phenotypes of controls and descendants (combined and children and grandchildren separately) were compared, taking into account multiple testing.

**Results:** Comparison of two generations descendants with corresponding control groups revealed significantly higher creatinine level and lower glomerular filtration rate (GFR), both in meta-analysis and in both generations independently. The mean values of GFR for all groups were detected within the normal range. Additionally, independent of the creatinine level, differences in the eating pattern were detected: insufficient fish consumption and excessive red meat consumption were significantly more frequent in the children of the Leningrad siege survivors compared with controls. BP and blood lipids did not differ between the groups.

**Conclusions:** Ancestral famine exposure in the prenatal period and early childhood may contribute to a decrease of in kidney filtration capacity and altered eating pattern in at least two generations of besieged Leningrad offspring.

## Introduction

The theory of fetal origin of health and diseases suggests that parental environmental exposure during fetal development may be an important risk factor for adult-onset diseases, in particular, for cardiovascular diseases (CVD) (1). Susceptibility to diseases resulting from fetal programming can be observed directly after birth in exposed individuals or transmitted from the exposed generation to the descendants.

Major famine studies of the Dutch Famine 1944-1945 and the Chinese Famine 1959-1962 have shown that insufficient nutrition during prenatal and early life periods have critical impacts on fetal programming. Importantly, not only were individuals directly exposed to famine during the prenatal period shown to have phenotypic consequences of exposure in later life. It was suggested that the effect of fetal programming can be transmitted to the descendants of exposed individuals, potentially through epigenetic mechanisms. For example, the Dutch Hunger study observed a higher frequency of neonatal obesity in children of women exposed to malnutrition prenatally and a higher body mass index (BMI) at an older age in the offspring of prenatally malnourished men (2,3). The China Health and Nutrition Survey reported significantly lower glomerular filtration rate (GFR), and a faster than expected increase in BMI with age, waist circumference, and blood pressure (BP) in the first generation of descendants of exposed individuals (4,5). Therefore, transgenerational transmission of the famine exposure phenotype could be observed across at least one generation of descendants.

The Siege of Leningrad was another tragic example of catastrophic malnutrition of the city’s residents having experienced a major supply deficit for two and a half years during the Second World War (6). There are some similarities but also difference with the two other major famine studies: The Siege of Leningrad was longer than Dutch Famine (28 months compared to about 6 months, respectively), the weather was significantly colder, affecting the calorie demand and potential hypothermia, and daily rations were not restored after breaking the blockade (in the Netherlands daily rations quickly rose within a week) (7). The Chinese famine, which is comparable in duration to the famine during the Siege of Leningrad, was characterized by pandemic nature and uneven intensity distribution within the country (8). And both previously mentioned studies involved only one generation of descendants.

The purpose of our study was to investigate the association of starvation in the prenatal period and the period of early childhood with phenotypic differences later in life in two generations of descendants of Leningrad Siege survivors (DLSS).

## Materials and methods

In 2009-2011, 305 Leningrad siege survivors who starved in early childhood and/or in utero during the Second World War (1941-1944) were examined in the Almazov National Medical Research Centre as part of an observational cohort study, the design and general characteristics were described in previous publications (6). In 2020-2021, we examined 87 adult DLSS (57 children and 30 grandchildren from 55 families) of 58 Siege survivors, with the current pregnancy being the only exclusion criteria (**Figure 1A**). A subsample of a population-based cohort (N=1,600) of St. Petersburg residents aged 25-64 years within the national epidemiological study ESSE-RF (9) were selected as controls by matching gender, age and BMI distribution to each generation of descendants. In the current study we refer to Leningrad Siege survivors exposed to starvation in early life as generation zero (F_0_), their children as the first generation of DLSS (F_1_) and grandchildren as the second generation (F_2_). The final control cohorts included 127 individuals for F_1_ and 48 for F_2_.

**Figure 1.**
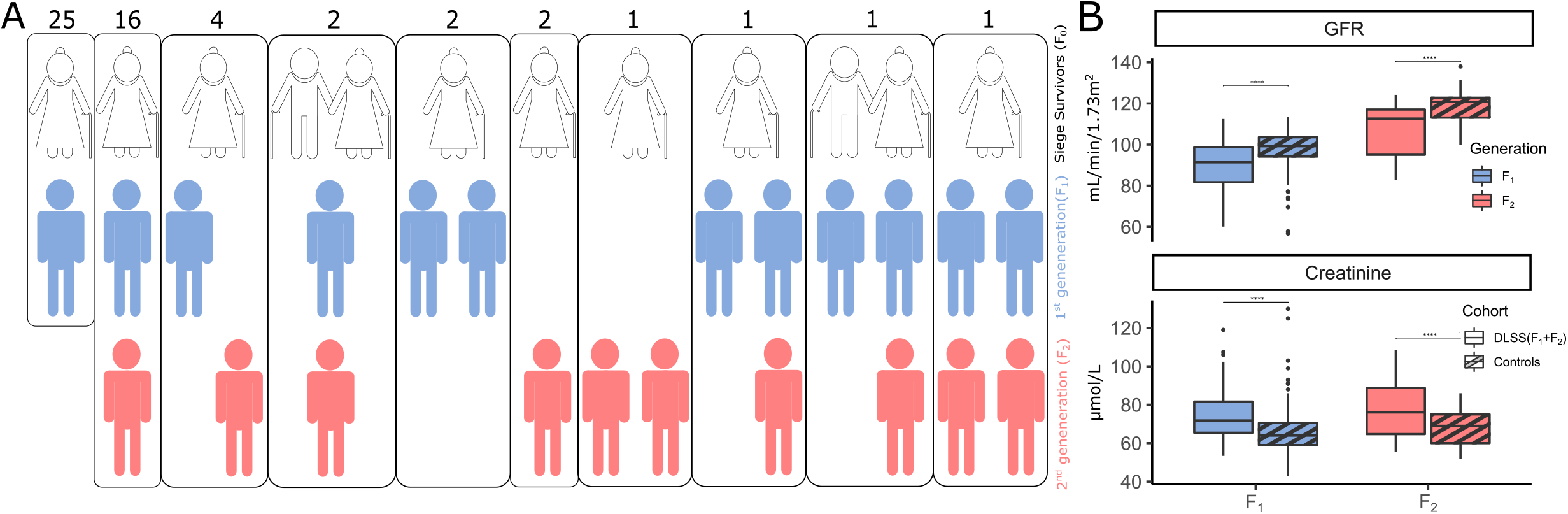
Comparison of descendants of Leningrad Siege survivors (DLSS) with population controls from St. Petersburg. **(A)** Graphical representation of all family trees in the DLSS cohort (Siege survivors -F_0_, the 1^st^ generation of descendants - F_1_, the 2^nd^ generation of descendants - F_2_); **(B)** Differences in GFR and creatinine levels between each generation of DLSS and the corresponding controls from the populational cohort of St. Petersburg.

The study was approved by the Local Ethics Committee of the Almazov National Medical Research Centre (Protocol #2401-21 from the local ethics committee meeting #01-21 dated January 18, 2021). All participants signed an informed consent.

Body weight measurement was carried out on the scales of the brand VEM-150-Mass-K (Russia), height - using the height meter RM-1 Diacoms (Russia), waist and hip circumference - using a standard flexible centimeter tape. Abdominal obesity was determined according to the JIS 2009 metabolic syndrome criteria 2009 (10) as: waist circumference ≥94 cm for males and ≥80 cm for females. The BMI was calculated as the ratio of body weight in kilograms to height in meters squared. All respondents were classified as obese (BMI ≥ 30 kg/m^2^) or non-obese (BMI <30 kg/m^2^).

Double measurement of blood pressure (BP) on the right arm was performed with the automatic tonometer “Omron” (Japan) with intervals of 2 minutes, after resting for 5 minutes in a sitting position, then a single measurement of BP was made 3 minutes after verticalization was made. Hypertension was diagnosed if the mean BP level was ≥140/90 mmHg and/or if patient was receiving antihypertensive therapy.

The creatinine, blood glucose, lipids study (total cholesterol – TC, low-density lipoproteins – LDL, high-density lipoproteins – HDL, triglycerides – TG) measurement was performed in fasting state (AbbotArchitect 8000 (USA)). The levels of leptin (Leptin ELISA, DBC (Canada)) and adiponectin (Adiponectin ELISA, Mediagnost (Germany)) levels were determined by enzyme immunoassay. We assessed glucose metabolism disorders: participants were divided into fasting hyperglycemia group (glucose level ≥5.6 mmol/L) and diabetes mellitus group (the patients self-reported about diabetes). The GFR was calculated using the CKD-EPI formula (11). Groups of participants with high total cholesterol (>4.9 mmol/L), LDL (>3.0 mmol/L), triglycerides (≥1.7 mmol/L), and reduced HDL (in male <1.0 and in female <1.2 mmol/L) were also formed; hypolipidemic therapy was taken into account (12).

Behavioral and socioeconomic risk factors were evaluated with questionnaires and interviews: diet pattern, smoking status, alcohol consumption, duration of sleep, level of education, physical activity. The consumption of fresh vegetables and fruits was assessed (respondents were divided into those who had fresh fruits and vegetables in their daily diet and those who consumed irregularly). Excess salt intake was considered to be salting of ready-made foods. Consumption of fish (200 grams) less than 1-2 times a week was considered insufficient. Daily consumption of red meat (150 grams or more) was considered excessive (13). If the respondent consumed ≥6 pieces/spoons of sugar per day or daily/almost daily intake of sweets/confectionery, it was considered excessive intake of sweets. The duration of sleep of less than 6 hours a day was considered insufficient. Moderate physical activity ≥150 minutes per week was sufficient. Inactivity was recorded in the case of being in a sitting position for more than 9 hours on a weekday. The assessment of smoking status was also carried out, respondents were divided into groups: smoking at the moment, in the past, and those who have never smoked. The number of alcohol doses and the frequency of alcohol consumption were assessed and respondents were divided into those who regularly took alcoholic beverages and those who did not consume at all.

Mathematical and statistical data analysis was implemented using the R-4.0 programming language (14) and libraries dplyr (v1.0.7) (15), ggplot2 (v3.3.5) (16), tidyr (v1.1.4) (17). Quantitative parameters were described using median values and lower and upper quartiles (Q1-Q3). Nominal data were described with absolute values and percentages.

The prevalence of 44 phenotypic risk factors in both generations of DLSS was simultaneously assessed using a logistic model adjusted for sex, generation and BMI. In case of absence of a phenotype in one of the comparison groups, a simple Fisher test was used instead of logistic regression. We report both effect size (beta) and odds ratio for each phenotype. If a phenotype was obtained from the other phenotype through calculations, we considered the pair as a single phenotype for multiple hypothesis correction with Bonferroni approach. In total, 34 independent phenotypes were analyzed and the significance threshold for the p-value was 0.05/34=0.0015.

## Results

87 DLSS (mean age: 43.8 years; sd: +/-13.4 years; range: 18-63 years; 44% men) were observed during an ambulatory visit at Almazov National Medical Research Centre (St. Petersburg, Russia). The prevalence of abdominal and obesity did not differ significantly among BMI-matched participants. The BP levels and the prevalence of hypertension was non-remarkable compared to population average. The DLSS group had nominally higher HDL levels. The differences are described in more details in the supplementary and Tab. S1.

The DLSS cohort had 57 and 30 individuals from the first and F_2_, respectively (**Fig. 1A**). Initially we performed matching of the DLSS cohort to a group of 1,600 controls from the ESSE population cohort from St. Petersburg. We excluded individuals with known cardiovascular diseases and outliers (more than 6 standard deviations) based on blood biochemical analyses and blood pressure measurements from the control pool. The final control pool available for matching had 1,142 individuals.

Controls of the same gender from the ESSE populational cohort were selected for each individual so that the age and BMI of the controls did not differ for more than two units. No controls were found for 8 DLSS individuals (7 youngest individuals from F_2_) and (1 individual with high BMI from F_2_), therefore they were excluded from further analyses. 3 individuals from F_1_ of DLSS were excluded because they had cardiovascular disease. The final cohort included a total of 175 controls and 76 DLSS (127 controls and 54 DLSS for F_1_; 48 controls and 22 DLSS for F_2_, **Sup. Fig. S1**).

Of all the biomarker phenotypes that were analyzed and compared, only creatinine (p=1.367×10^−7^, beta=0.092, se=0.017) and GFR (p=3.94×10^−8^, beta=-0,08, se=0.015) passed the Bonferroni significance threshold (**Fig. 1B, Sup. Tab. S1**). Additionally, insufficient fish consumption was more frequently observed in the DLSS group compared to controls (p = 3.15×10^−6^ ; beta = 1.361; beta se=0.292), and also excessive red meat consumption was more frequently observed in the DLSS group (p = 1.8×10^−4^; beta=1.168 ; beta se=0.312) (Sup. Tab. S1). Among cardiometabolic risk factors, nominal differences were observed for leptin and HDL levels that were higher in the DLSS group, but still within normal ranges (leptin: p = 3.12×10^−3^, beta=0.037; HDL: p = 9.47×10^−3^, beta=1.08) and not significant after multiple hypothesis testing correction.

Further, we investigated whether this effect could be observed in each generation separately. We analyzed the phenotypic data within each generation independently, adjusting the model for age, BMI, sex. Interestingly, the effect of creatinine and GFR was observed in both generations - F_1_: creatinine (p=9.71×10^−5^, beta=0.071, se=0.018) and GFR (p=1.09×10^−5^, beta=-0.076, se=0.017) and F_2_: creatinine (p=3.56×10^−4^, beta=0.208, se=0.058) and GFR (p=4.56×10^−4^, beta=-0.179, se=0.051) (Sup. Tab. S1). The differences in fish and red meat intake habits passed Bonferroni correction only for F_1_ (red meat: p = 3.10×10^−4^; beta=1.39; beta se=0.385. Fish: p = 5.23×10^−4^; beta = 1.197; beta se=0.345), but estimates for F2 were in similar direction and magnitude (red meat: p = 0,237; beta = 0,653; beta se = 0.55. Fish: p = 0,0015; beta = 1,897; beta se = 0.598) (Sup. Tab. S1).

In addition to the consumption of red meat and fish, several other nutritional phenotypes were nominally significant, therefore, we used the principal component analysis (PCA) based on food consumption-related phenotypes (fish, red meat, salt, sugar, vegetable intake, and leptin) to investigate differences in food pattern between DLSS and controls (**Sup. Fig. S2A**). We found that PC2, which was characterized by a low fish intake, leptin levels, and high salt and sugar intake, and differed significantly between descendants and controls for both the meta-analysis (p = 3.77×10^−9^; beta=-1.06 ; beta se=0.18) and for each generation separately: F_1_ (p = 2.15×10^−6^; beta=-1.00 ; beta se=0.211); F_2_ (p = 5.29×10^−4^; beta=-1.243; beta se=0.359) (**Sup. Tab. S2**).

Subsequently, we investigated whether the food pattern was related to the observed difference in creatinine level. From the control population that was not used in the previous comparisons, we selected individuals to match sex, age, and PC2 distributions obtained from the PCA of food intake in the DLSS cohort (**Sup. Fig. S2B**). Comparison of this cohort with the previously selected control cohort for DLSS comparison, we did not observe significant differences in creatinine and GFR (p=0.75; p=0.61, respectively, **Sup. Fig. S2C**). Therefore, the pattern of food consumption and biochemical markers of renal function can be interpreted as independent associations.

## Discussion

Our results demonstrate a significantly higher creatinine level and a lower glomerular filtration rate in the descendant generations of individuals exposed to prenatal or early childhood starvation. It is known that most human organs have a critical development period at the prenatal stage. In the case of serious nutritional deficiency, the number of functioning glomeruli decreases, leading to increased filtration load, increased GFR throughout postnatal life, and subsequent damage to functioning glomeruli (18). Reports on the Northern China famine cohort study also presented significantly lower calculated GFR in prenatally exposed participants to starvation and their children compared to controls. Data on the second generation were not available in this study. It has been hypothesized that the starvation of the ancestors in early childhood contributes to a decrease in the renal filtration capacity in their offspring. This may be due to transgenerational changes in the epigenome through the programming of nephron deficiency in germ cells (4). In the long term, renal filtration capacity can cause glomerulosclerosis, and finally hypertension in adulthood (18).

Observed differences in the dietary pattern between the descendants and controls do not have a clear direction of effect. On the one hand, increased red meat and decreased fish consumption increase the risk of cardiometabolic disorders (19,20). On the other hand, the decreased salt consumption and higher leptin levels might indicate a more healthy approach to food selection. Comparison of the subsample of the population cohort that matched in dietary behavior with DLSS with the control population indicated that the observed differences in creatine levels and food selection were independent of each other. Therefore, it is likely that there is an independent effect of dietary behavior imprinting observed in DLSS.

The absence of statistically significant differences in the prevalence of cardiometabolic diseases can be explained on the one hand by the small size of the cohort studied, and on the other hand by the relatively young age of most of the participants. Also, it is important to note that in this work we studied the offspring, among whom most of the parents are still alive, which may have resulted in survivorship bias. In addition, both a decrease in fertility and an increase in perinatal mortality during famine may have led to selective births of healthier or more resilient individuals. The besieged Leningrad residents included in the study could therefore represent an artificially “healthy” sample.

Conclusively, observation of phenotypic effects associated with early-in-life exposures, separated not only in time, but also by generations serve yet another example of Barker’s hypothesis of fetal origins of chronic diseases of adult life. Our study suggests that not only individuals directly exposed to traumatic events, but their offspring will benefit from additional health screenings, in particular assessment of kidney function, for earlier detection of disease symptoms and their successful treatment.

## Data Availability

Raw data produced in the present study constitutes sensitive personal information and is not permitted for sharing by patient consent terms. Summary data characterizations are contained in the manuscript. More detailed information is available upon reasonable request to the authors.

## Author Contributions

K.T., D.U., O.R., M.A. designed and conceived the study.

K.T., D.U., E.M., M.B., E.K., A.E., T.V., O.R., M.A. collected and analyzed the data.

K.T., D.U., O.R., M.A. designed statistical analysis approach.

K.T., E.M., M.B., E.K., A.E., E.V., An.K., Al.K., O.R. recruited and supervised patients.

Al.K., E.S., M.A. – obtained funding.

O.R., M.A. supervised the study

K.T., D.U., O.R., M.A. wrote the manuscript

All authors edited and reviewed the manuscript text

