## Supplementary Materials for "TRANSGENERATIONAL EFFECT OF EARLY CHILDHOOD FAMINE EXPOSURE IN THE COHORT OF LENINGRAD SIEGE SURVIVORS’ OFFSPRING"

* - contributed equally

**The authors declare no conflict of interest**

**Table of contents**

**Controls Matching** **3**

**Phenotypic analysis** 4**Exploration of phenotypic patterns**

5

### Controls Matching

We used R to explore 87 descendants of Leningrad Siege survivors (DLSS) cohort and group of 1,600 control individuals from ESSE populational cohort (**Sup. Fig. 1A)** (1)**.**

Initially, we excluded all patients with cardio-vascular diseases and patients with more than 6 standard deviations of general biochemical parameters and blood pressure. Only 1,142 individuals were rested as potential controls.

Controls of the same gender from the populational cohort were selected for each individual so that the age and BMI of the controls did not differ more than 2. No controls were found for 8 DLSS individuals (7 young individuals from $F_2$) and (1 individual with high BMI from $F_2$).3 individuals from $F_1$ were excluded because they had cardiovascular disease. Finally, we found 175 controls for 75 cases (127/54 for the first population and 48/22 for the second population). (**Sup. Fig. 1B)**

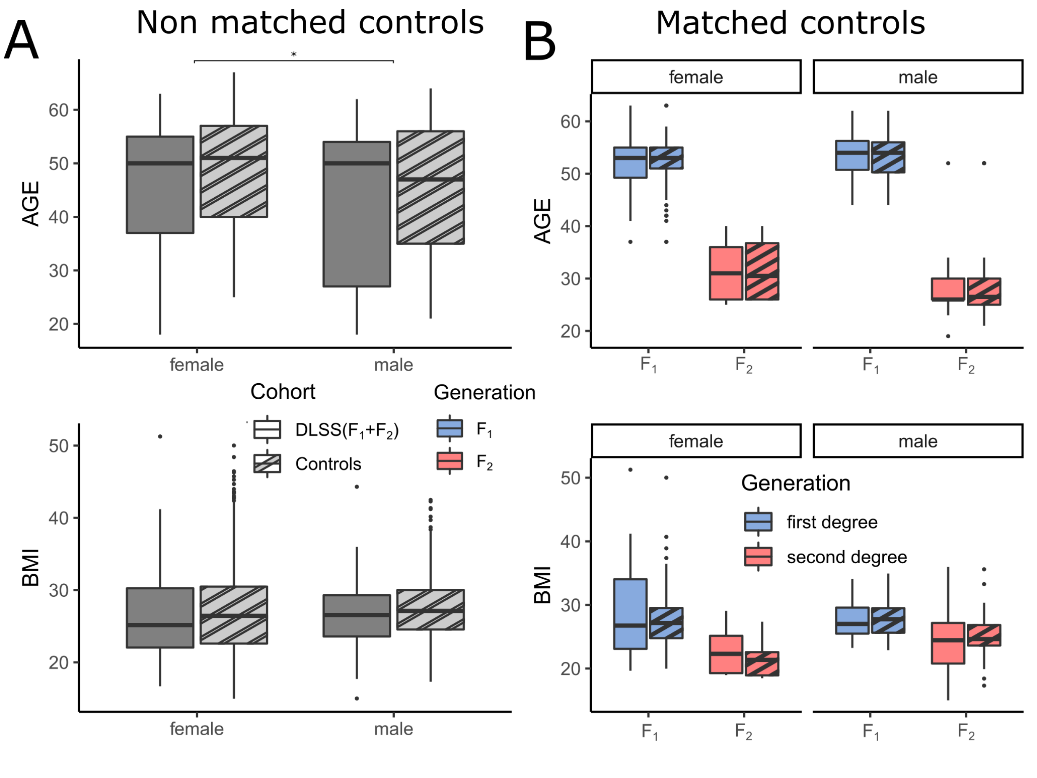


**Supplementary figure S1**. Comparison of DLSS with populational cohort **(A)** Non-matched controls; **(B)** Matched controls

### Phenotypic analysis

We used logistic regression model adjusted for sex generation and BMI to find differences of 44 phenotypic risk factors between DLSS and control groups. If a phenotype was obtained from the other phenotype through calculations, we considered the pair as a single phenotype for multiple hypothesis correction with Bonferroni approach - in total, 34 independent phenotypes were analyzed, therefore significance threshold was defined as 0.05/34=0.0015. Additionally, we implemented these models for each generation independently, replacing the generation correction in model with age.

Only creatinine ($p=1.367x10^{-7}$, beta=0.092, se=0.017), and GFR ($p=3.94x10^{-8}$, beta=-0,08, se=0.015) passed the significance threshold (Fig. 1B**, Sup. Tab. S1**). Additionally, insufficient fish consumption was more frequently observed in the DLSS group compared to controls ($p = 3.15x10^{-6}$; beta = 1.361; beta se=0.292) and excessive red meat consumption were more frequently observed in the DLSS group ($p = 1.8x10^{-4}$; beta=1.168; beta se=0.312) (Sup. Tab. S1)**.**

Also, we detected nominal differences in leptin and HDL levels that were higher in the DLSS group within normal ranges (leptine: $p = 3.12x10^{-3}$; beta=0.037, HDL: $p = 9.47x10^{-3}$; beta=1.08). Also, we found the less excessive salt consumption among DLSS group and more frequent diabetes mellitus (Salt: p=0.0056, beta=-0.814, diabetes: p=0.0068, beta =2.327)

Our previous research demonstrated a higher level of HDL within the normal range in besieged Leningrad residents compared to controls (2). Also, maternal nutrition during pregnancy also plays a key role in the proopiomelanocortin neurons development (satiety key mediator). The proopiomelanocortin neurons of the hypothalamus arched nucleus integrate peripheral signals such as leptin, glucose and insulin, and regulate the energy balance, causing a feeling of satiety and increasing energy consumption (3).

Attitude to low salt intake is one of the demonstrated by DLSS favorable behavioral patterns - they consumed an excessive amount of salt less often with no difference between two generations. According to V.S. Volkov, during the Siege of Leningrad, excessive salt intake was observed and was linked to high mortality due to hypertensive complications (4). Lower salt intake in the Siege of Leningrad survivors might be one of the favorable mechanisms of survival which was passed on to the offspring. Based on our biochemical data, it can be assumed that the descendants of the inhabitants of Besieged Leningrad more often have metabolic disorders, but some favorable behavioral factors may be helpful for preventing the disease's development. Thus, DLSS diet habits can have positive and negative effects on the health of study participants themselves and future generations through epigenetic changes. However, to assess long-term results, a longer follow-up of this cohort is needed.

**Investigation of phenotypic patterns.**

Since some phenotypes were of nominal significance we combined them into groups (phenotype codes are explained in Sup. Tab. S1): food pattern: HSALT, LFISH, HSUG, LFVI, M2_51_norm, leptin;

biochemical pattern: crea, GFR, adiponectin, chol, Tg, ldlp, hdlp, gluc;

disease pattern: SBP, DBP, AH, M8_6, grCHOL2, grTG2, grLDL2, grHDL2, M8_38, M8_39, AO_94_80, HyperGLU_56new, DIS17;

behavioral pattern: M3_2, SITHIGH, ALC, M4_1a, M4_1b, M4_1c, M7_1, M7_1_dop

For each pattern we independently performed PCA and extracted the first two principal components. Then for each PC we performed logistic regression with the model previously used for phenotypic analysis. We found that PC2 was significantly different (meta-analysis with two generations of descendants: $p = 3.77x10^{-9}$; beta=-1.06; beta se=0.18); ($F_1$: $p = 2.15x10^{-6}$; beta=-1.00; beta se=0.211); ($F_2$: $p = 5.29x10^{-4}$; beta=-1.243; beta se=0.359) (**Sup. Fig. S2A,** Sup. Tab. S1).

We hypothesized that food patterns could be related to the observed difference in creatinine levels. From a control population that was not used in previous comparisons, individuals were selected to match the sex, age, and PC2 distribution derived from the PCA of food intake in the DLSS cohort (**Sup. Fig. S2B**). When comparing this cohort with the previously selected DLSS control cohort, we did not observe significant differences in creatinine and GFR (p=0.75; p=0.61, respectively, **Sup. Fig. S2C**).

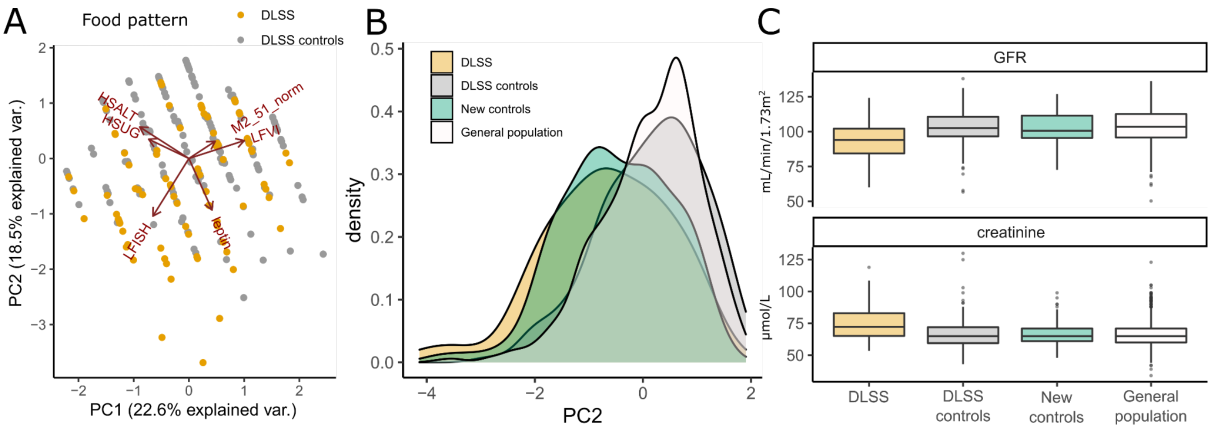


**Supplementary figure S2**. Food consumption pattern. **(A)** The first two principal component (PC) of food consumption pattern and loadings; **(B)** The distribution of PC2 in different cohorts; **(C)** The distribution of GFR and creatinine levels in different cohorts
